# Functional Coupling and Longitudinal Outcome Prediction in First-Episode Psychosis

**DOI:** 10.1101/2025.04.01.25325005

**Authors:** Isaac Z. Pope, Sidhant Chopra, Alexander Holmes, Shona M. Francey, Brian O’Donoghue, Vanessa L. Cropley, Barnaby Nelson, Hok Pan Yuen, Kelly Allott, Mario Alvarez-Jimenez, Susy Harrigan, Christos Pantelis, Stephen J. Wood, Patrick D. McGorry, Alex Fornito

## Abstract

**Background:** Clinical outcomesfollowing a first episode of psychosis (FEP) are highly heterogeneous between patients. The identification of prognostic biomarkers would greatly facilitate personalized treatments. Psychosis patients often display brain-wide disruptions of inter-regional functional coupling (FC), with some being linked to symptom severity and remission. FC may thus hold prognostic potential for people experiencing psychosis.

**Methods:** Ninety antipsychotic-naïve FEP patients (51% female, 15-25 years) were randomized to receive either antipsychotic or placebo tablets for 6 months alongside psychosocial interventions. A subset of these patients also completed functional magnetic resonance imaging (55 with usable data at baseline and 37 at 3 months), which was used to evaluate whether baseline FC, or 3-month change in FC, could predict 6- and 12-month changes in symptoms and functioning, quantified using the Brief Psychiatric Rating Scale and the Social and Occupational Functioning Assessment Scale, respectively. We considered three different cross-validated prediction algorithms: (i) connectome-based predictive modelling; (ii) kernel ridge regression; and (iii) multilayer meta-matching.

**Results:** All algorithms showed poor performance in predicting patients’ 6- and 12-month changes in symptoms and functioning (all *r*_*mean*_ < 0.3), and no model achieved significance via permutation testing (all *p* > 0.05).

**Conclusions:** Our findings suggest that brain-wide measures of FC may not be suitable for predicting extended clinical outcomes over a 6- to 12-month period in FEP patients.

## Introduction

Clinical outcomes following a first episode of psychosis (FEP) are heterogenous in both symptoms and functioning, ranging from permanent recovery to chronic and severe illness (1–4). While early intervention with pharmacological and psychosocial treatments has been shown to improve clinical outcomes in FEP patients (5–10), the efficacy and tolerability of these treatments vary considerably between individuals (11,12). Moreover, some FEP patients may recover through psychosocial treatment alone (13). Current clinical guidelines for FEP address this heterogeneity via trial-and-error in prescription (14,15), potentially delaying remission and recovery (16). The identification of reliable predictors of patients’ clinical outcomes is therefore a necessary step towards the development of personalized treatment strategies.

Across the psychosis spectrum, magnetic resonance imaging (MRI) studies have revealed disruptions in both the structural connectivity between different brain regions (17–19), and the patterns of inter-regional functional coupling (FC; i.e., correlated activity) that these connections support (18,20–28). Some of these FC disruptions are apparent at the onset of psychosis (20–22,27,28) and in high risk individuals (29) and are related to different symptoms (29–31). Most importantly, prospective studies have found associations between patients’ baseline FC patterns and their changes in symptoms and functioning following treatment (28,32–36). Despite the promise of this work, the analyses have generally relied on within-sample quantification of associations between FC and outcomes, which may inflate effect size estimates due to over-fitting (37). A thorough assessment of the generalizability of any putative prognostic biomarker therefore requires analytic strategies such as cross-validation (37).

Some studies have reported cross-validated evidence that baseline FC can predict longitudinal symptom changes in FEP patients (38–44), but most only involved short-term outcomes (i.e., up to 16 weeks post-baseline) in patients diagnosed with schizophrenia, potentially impedingthe generalizability of their findings to the kindsof transdiagnostic FEP cohorts encountered in real-world clinical care (45). Moreover, a focus on predicting symptom changes may not translate to an impact on functional outcomes, since the two are not always related (46,47). Functional recovery in domains such as social relationships, educational attainment, and occupational functioning is particularly important in this regard, as it may better address patient needs compared to symptom recovery (48). Critically, all prior studies only use a single prediction algorithm, rather than comparing the performance of multiple gold-standard approaches, limiting our ability to establish reliable and reproducible prediction methods for FEP.

In this study, we apply multiple cross-validated prediction algorithms to task-free resting-state functional MRI (fMRI) and clinical outcome datafrom the Staged Treatment and Acceptability Guidelines in Early Psychosis Study (STAGES), a triple-blind randomized control trial of antipsychotics in previously antipsychotic-naïve FEP patients (13,49). Our aim was to assess whether patients’ baseline resting-state FC could predict their changes in symptoms and functioning after 6 and 12 months. We secondarily evaluated whether longitudinal changes in FC between baseline and 3 months (ΔFC) could predict the same outcomes.

## Methods and Materials

### Design and Participants

This study used clinical and neuroimaging data from STAGES (13,49), which was approved by the Melbourne Health Human Research and Ethics Committee and registered under the Australian New Zealand Clinical Trials Registry (ACTRN12607000608460) in November 2007. Ninety FEP patients (ages 15-25 years) were recruited between 2008 and 2016 at the Early Psychosis Prevention and Intervention Centre at Orygen Youth Health in Melbourne, Australia. Eligibility requirements comprised: (i) meeting criteria for a psychotic disorder in the DSM-IV via a Structured Clinical Interview; (ii) a duration of untreated psychosis under 6 months; (iii) ability to provide informed consent; (iv) low risk of suicidality, self-harm, or hostility; (v) negligible lifetime use of antipsychotics, or present use of mood stabilizers; (vi) stable housing and support; and (vii) no pregnancy. Patients were randomized to receive antipsychotic medication (risperidone or paliperidone) or matched placebo tablets for 6 months, along with psychosocial intervention in the form of cognitive behavioral case management (50). Further details about the trial design, including safety protocols, can be found in the supplement and previous publications (13,49).

Patients’ functioning and symptoms were measured at baseline and after 6 and 12 months via the Social and Occupational Functioning Assessment Scale (SOFAS) and the Brief Psychiatric Rating Scale (BPRS), respectively (Figure 1). Patients’ total scores on these inventories were preregistered as the primary and secondary outcomes of the larger clinical trial, respectively. Since we have previously demonstrated no differences in these outcomes between the placebo and medicated patients at 6 or 12 months, both treatment groups were combined in predictive models to maximize statistical power and generalizability. Since dichotomizing of patients into ‘responders’ and ‘non-responders’ is statistically inefficient in ignoring within-group variability (51), and is not supported by a consensus definition of clinical recovery (3), we defined patients’ clinical outcomes as continuous proportional change scores (*y*_2_ − *y*_1_)/*y*_1_, where *y*_1_ and *y*_2_ denote their SOFAS or BPRS score at baseline and at a later timepoint (6 or 12 months), respectively. Despite attrition throughout the trial, most patients who discontinued the trial still chose to complete clinical assessments. We included their data in our analysis to maximize statistical power and generalizability.

**Figure 1:**
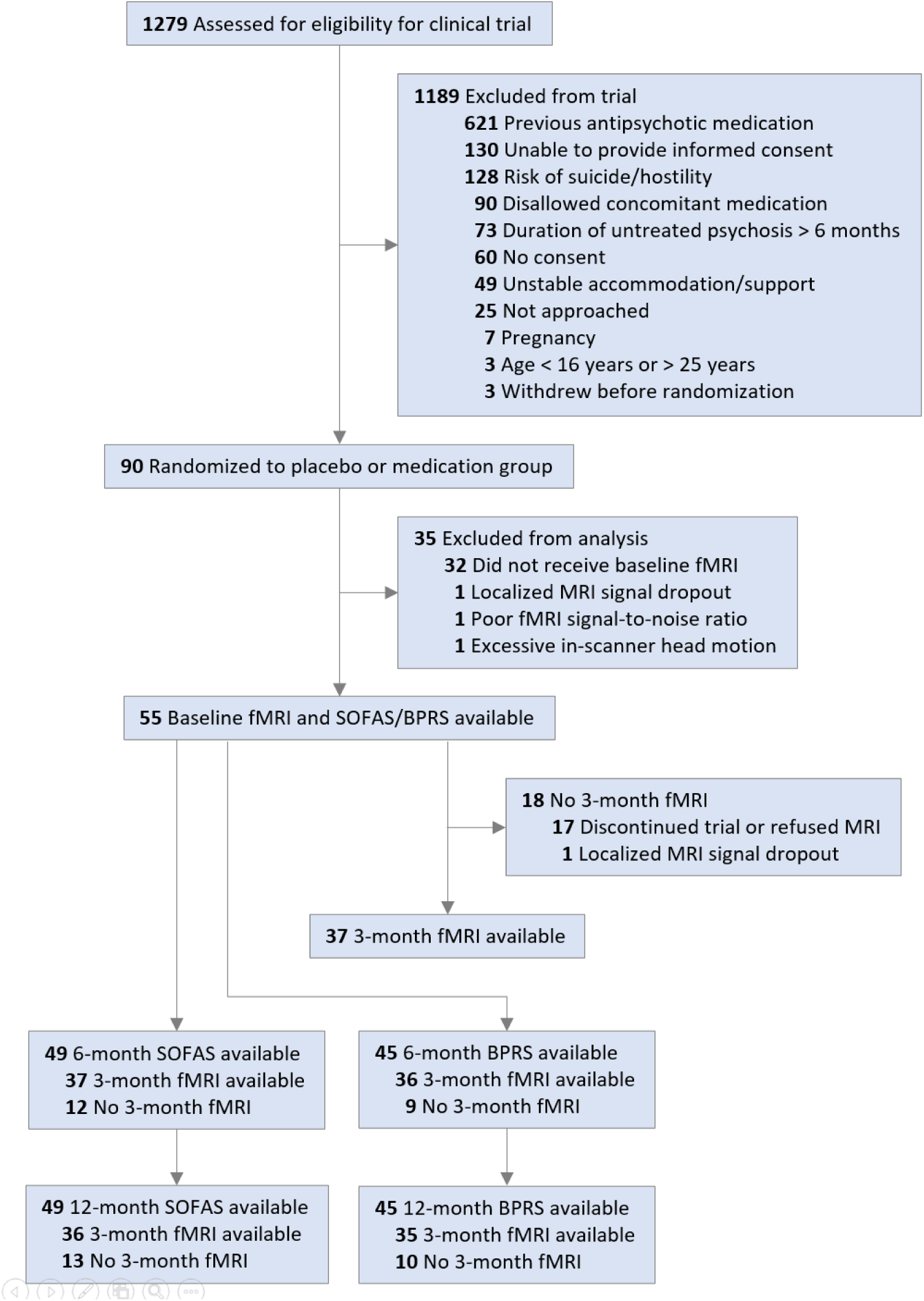
Flow diagram for patients involved in the larger clinical trial and in the present analysis. MRI, magnetic resonance imaging; fMRI, functional MRI; SOFAS, Social and Occupational Functioning Assessment Scale; BPRS, Brief Psychiatric Rating Scale.

### MRI Acquisition and Processing

A 3T Siemens Trio Tim scanner with a 32-channel head coil was used to acquire T1-weighted anatomical MRI and resting-state fMRI data at the Royal Children’s Hospital in Melbourne, Australia. Of the 90 patients in the clinical trial, 58 and 39 were scanned at baseline and after 3 months, respectively. We used the software MRI-QC to automatically compute imagequality metricsforeach scan(52), then excluded three scans fromfurther analysisdue to localized signal dropout or high noise. We also removed one scan due to excessive head motion, classified using standard exclusion criteria (53). The remaining scans were preprocessed via fMRIPrep v1.4.1 (54), then denoised using a pipeline previously shown to mitigate the impacts of noise sources such as head motion and non-neuronal physiological fluctuations (53). For each voxel in a scan, this involved linear detrending, regressing out motion-related signals automatically identified by the software ICA-AROMA (55), regressing out mean signals of grey matter, white matter, and cerebrospinal fluid tissues, and band-pass temporal filtering at 0.008-0.08 Hz (Figure S1). Each brain was parcellated using the 300-region Schaefer cortical atlas (56) and the Melbourne Scale II subcortical atlas (57), producing a mean fMRI signal time series for each region. We then computed Pearson’s correlation for every pair of these time series to generate a 328 × 328 FC matrix for each scan.

Since one prediction algorithm that we used (multilayer meta-matching) has been pre-trained on FC from larger datasets, its use requires that FC input shares the same parcellation as its training data (58). Accordingly, we combined the 400-region Schaefer cortical atlas with the automated subcortical segmentation in FreeSurfer (59,60) to separately produce the required 419 × 419 FC matrices.

For ΔFC-based predictions, matrices were instead derived by subtracting each patient’s baseline FC matrix from their 3-month FC matrix. This means that each entry in a patient’s ΔFC matrix contains their longitudinal change in FC over the first 3 months, for some coupling of brain regions. Further details on acquisition parameters, quality control, and preprocessing are provided in the Supplement.

### Predictive Modelling

To evaluate the robustness of our findings, we predicted patient outcomes using three different algorithms: (i) connectome-based predictive modelling (CPM); (ii) kernel ridge regression (KRR); and (iii) multilayer meta-matching. The first two are commonly used in the field. The last has been recently shown to improve the performance of predictive models in small samples (58). All algorithms performed 4-fold cross-validation, where data are first divided into four equally-sized folds.Three folds are used for training a model while the remaining fold is held asidefor testing this model via prediction. This procedureis repeated four times, with each fold used for testing once. Thefollowing presents details of each prediction algorithm.

#### Connectome-based Predictive Modelling

CPM is a simple approach that prioritizes direct interpretability of brain regions and networks implicated in accurate predictions (61). We implemented CPM via 16 models, which differed in terms of the: (i) outcome measure (symptoms or functioning); (ii) outcome timepoint (6- or 12-month change); (iii) predictor (baseline FC or ΔFC); and (iv) feature selection (positive or negative). In each model, CPM first performs feature selection in the training set, using Pearson’s correlation to identify FC estimates that are either positively (*r* > 0, *p* < 0.05 uncorrected) or negatively(*r* < 0, *p* < 0.05 uncorrected) associated with outcomes. For each patient, FC values are then summed across these positive or negative features to create a summary score. A linear model is fitted between these FC summary scores and outcomes within the training set, then used to predict the outcomes of patients in the testing set.

#### Kernel Ridge Regression

KRR is a classical machine learning method that lacks a feature selection step, instead performing multivariable prediction with regularization, where a hyperparameter is optimized to control the trade-off between training error and testing error (62). We used the resting-state FC-based KRR implementation from Li et al. (2019) (63), which has been shown to predict many behavioral measures with accuracy comparable to more computationally demanding deep neural networks (64). KRR was implemented via eight models, which differed in terms of the: (i) outcome measure (symptoms or functioning); (ii) outcome timepoint (6- or 12-month change); and (iii) predictor (baseline FC or ΔFC). In KRR, each outcome in the testing set is predicted as a weighted mean of the outcomes observed in the training set. These weights include measures of similarity in FC between patientsand an*l*_2_-regularization hyperparameter whosevalue is optimized via an inner loop of 4-fold cross-validation within the training set (further details are provided in the Supplement).

#### Multilayer Meta-matching

Recent work suggests that prediction models that perform well within a small sample often fail to generalize across datasets (65–69). To boost our prediction accuracies while avoiding overoptimism, we additionally used multilayer meta-matching, a transfer learning framework that transposes prediction models trained on large healthy datasets to a smaller dataset of interest. This approach exploits neural degeneracy, whereby a relatively small set of resting-state FC patterns underlie many phenotypes spanning cognition, demographics, and mental health (70–72). We used the pre-trained model described in Chen et al. (2024), which significantly outperformed classical KRR in samples as small as 10 individuals (58) and significantly predicted cognitive measures within and across multiple psychiatric cohorts (73).

Briefly, this involved using a deep neural network and linear ridge regression to predict 67 different phenotypes in the UK Biobank dataset (*n* = 36834) (74) based on resting-state FC. These techniques were then used to predict another 162 phenotypes in other datasets (Adolescent Brain Cognitive Development, *n* = 5985 (75); Genomics Superstruct Project, *n* = 862 (76); Healthy Brain Network, *n* = 930 (77); Enhanced Nathan Kline Institute Rockland Sample, *n* = 896 (78)). We used this pre-trained model to predict all phenotypes for each STAGES patient, thereby generating 458 proxy variables that replaced FC as the inputs for a final KRR step to predict the clinical outcomes. Since no models have yet been pre-trained using longitudinal changes in FC, we implemented multilayer meta-matchingvia four models, which differed in terms of: (i) outcome measure (symptoms or functioning); and (ii) outcome timepoint (6- or 12-month change).

### Evaluating Prediction Performance and Significance

To further reduce sampling bias, we ran each of the above models across 100 splits, where each split used a different random allocation of patients to the four folds. We then quantified each model’s prediction performance as the mean correlation between predicted and observed outcomes across all 100 splits (*r*_*mean*_). We assessed prediction significance via permutation testing, which generates empirical null distributionsof *r*_*mean*_. For each model, outcomes were randomly shuffled amongst patients 1000 times, then prediction algorithms were rerun for each of these permutations. 100 splits per permutation were used for CPM null models, but to reduce computational burden we used 50 splits for KRR and 20 splits for multilayer meta-matching. We calculated *p*-values as the proportion of null *r*_*mean*_ values that exceeded the true *r*_*mean*_.

To correct for family-wise error (FWE) arising from the inclusion of multiple predictive models, the inference-based Westfall-Young method was chosen (79). For this, the highest *r*_*mean*_ across all 16 null CPM models was retained at each of the 1000 permutations, producing a single FWE-corrected null distribution for the calculation of all 16 *p*_*FWE*_-values. FWE correction was similarly applied across all eight KRR models and across all four multilayer meta-matching models. This ensures that models only achieve significance by surpassing the strongest prediction performances across all null models sharingthe same prediction algorithm. We chose this approach to reduce false positives in a moretailored manner compared to alternative methods such as Bonferroni correction, which may have inflated the rate of false negatives (80,81). Statistical significance was assessed at *p*_*FWE*_ < 0.05.

## Results

### Clinical Outcomes

Across all patients included in our analysis, 73% had increased functioning (SOFAS) after the 6 months of STAGES treatment (mean proportional change = 20%, *n* = 49; Figure S2) and 69% had increased after 12 months (mean proportional change = 22%, *n* = 49). Similarly, 84% of patients had decreased symptoms (BPRS) after 6 months (mean proportional change = −25%, *n* = 45) and 84% had decreased after 12 months (mean proportional change = −23%, *n* = 45). Patients’ baseline SOFAS scores (52.3 [12.1]; mean [SD]; Table S1) were comparable to those of a large epidemiologically representative FEP cohort (82), and baseline BPRS total scores (57.6 [9.4]) indicated marked illness (83). Patients who discontinued prior to 3 months did not receive a second MRI scan, meaning predictive models based on ΔFC used subsamples of 35-37 patients. Clinical outcomes did not significantly differ between patients with 3-month FC data and those without (two-tailed *t*-test, *p* > 0.05 for all 6- and 12-month SOFAS and BPRS data).

### Predictive Modelling

#### Connectome-based Predictive Modelling

The eight CPM models that used baseline FC to predict outcomes showed limited predictive value (*r*_*mean*_ range of −0.27 to 0.17; Table 1 and Figure 2). This was also the case for the eight ΔFC models (*r*_*mean*_range of −0.28 to 0.18). Permutation testing via empirical null distributions revealed that none of the CPM models passed the threshold for statistical significance before or after FWE correction (all *p* ≥ 0.07; Table 1 and Figure S3). We found similar results when using alternative feature selection thresholds of *p* < 0.05 and *p* < 0.001 in CPM (all *r*_*mean*_ < 0.3; Table S2-S3, Figures S6 and S8) or significance (all *p* ≥ 0.06; Table S2-S3, Figures S7 and S9).

**Figure 2:**
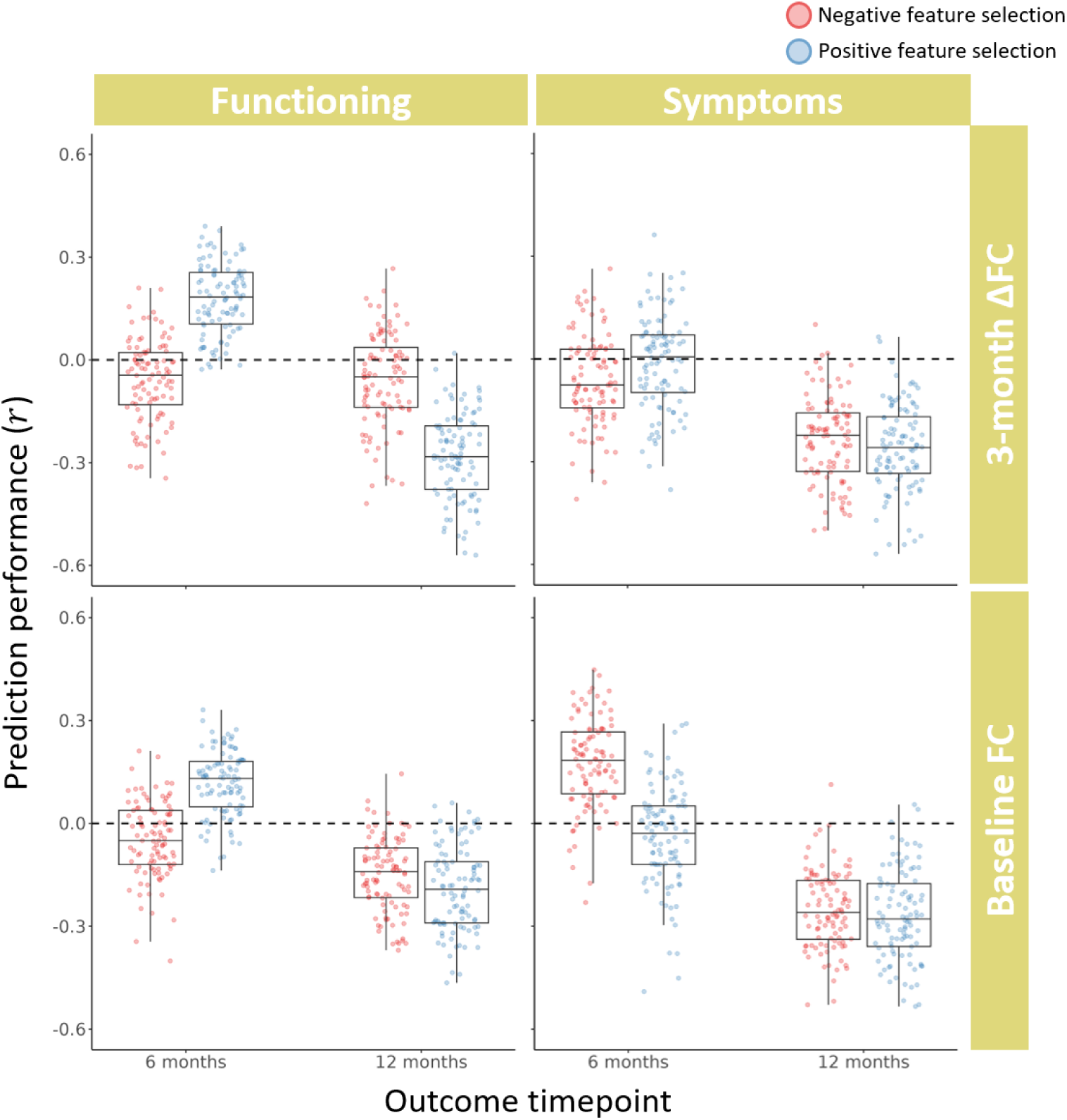
Performance of connectome-based predictive modelling (CPM) for predicting patients’ 6- and 12-month changes in symptoms and functioning, using either baseline functional coupling (FC) or 3-month change in FC. Each data point shows the strength of Pearson’s correlation between predicted and observed clinical outcomes for a single split of 4-fold cross-validation, with each of the 16 models comprising 100 random splits.

**Table 1:**
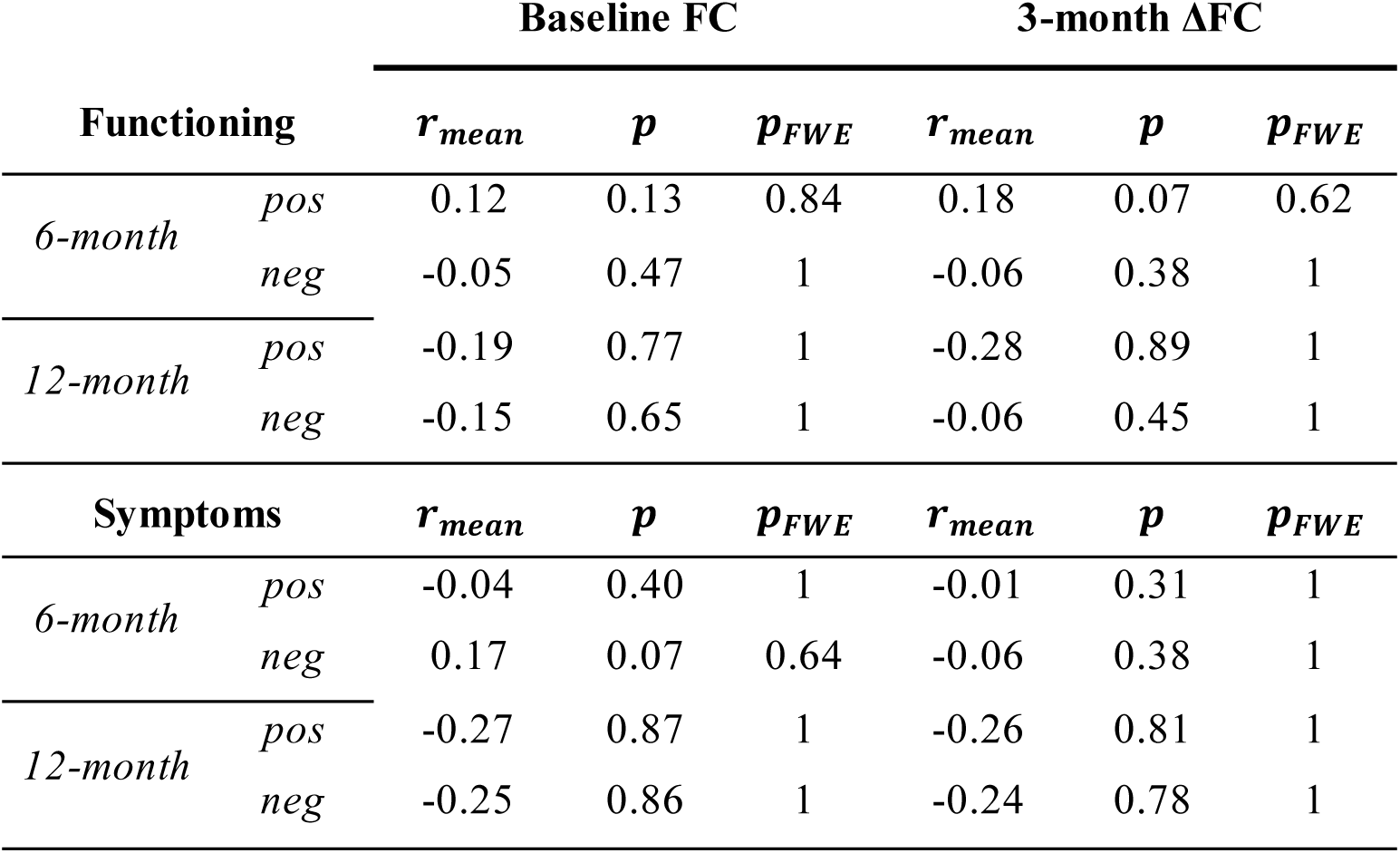
Prediction performance and significance for all 16 connectome-based predictive modelling (CPM) models. pos, positive feature model; neg, negative feature model.

#### Kernel Ridge Regression

The four KRR models that used baseline FC to predict outcomes also showed limited predictive value (*r*_*mean*_ range of −0.24 to 0.23; Table 2 and Figure 3). This was also the case for the four ΔFC models (*r*_*mean*_range of −0.28 to 0.21). Permutation testing via empirical null distributions revealed that none of the KRR models passed the threshold for statistical significance before or after FWE correction (all *p* ≥ 0.12; Table 2 and Figure S4).

**Figure 3:**
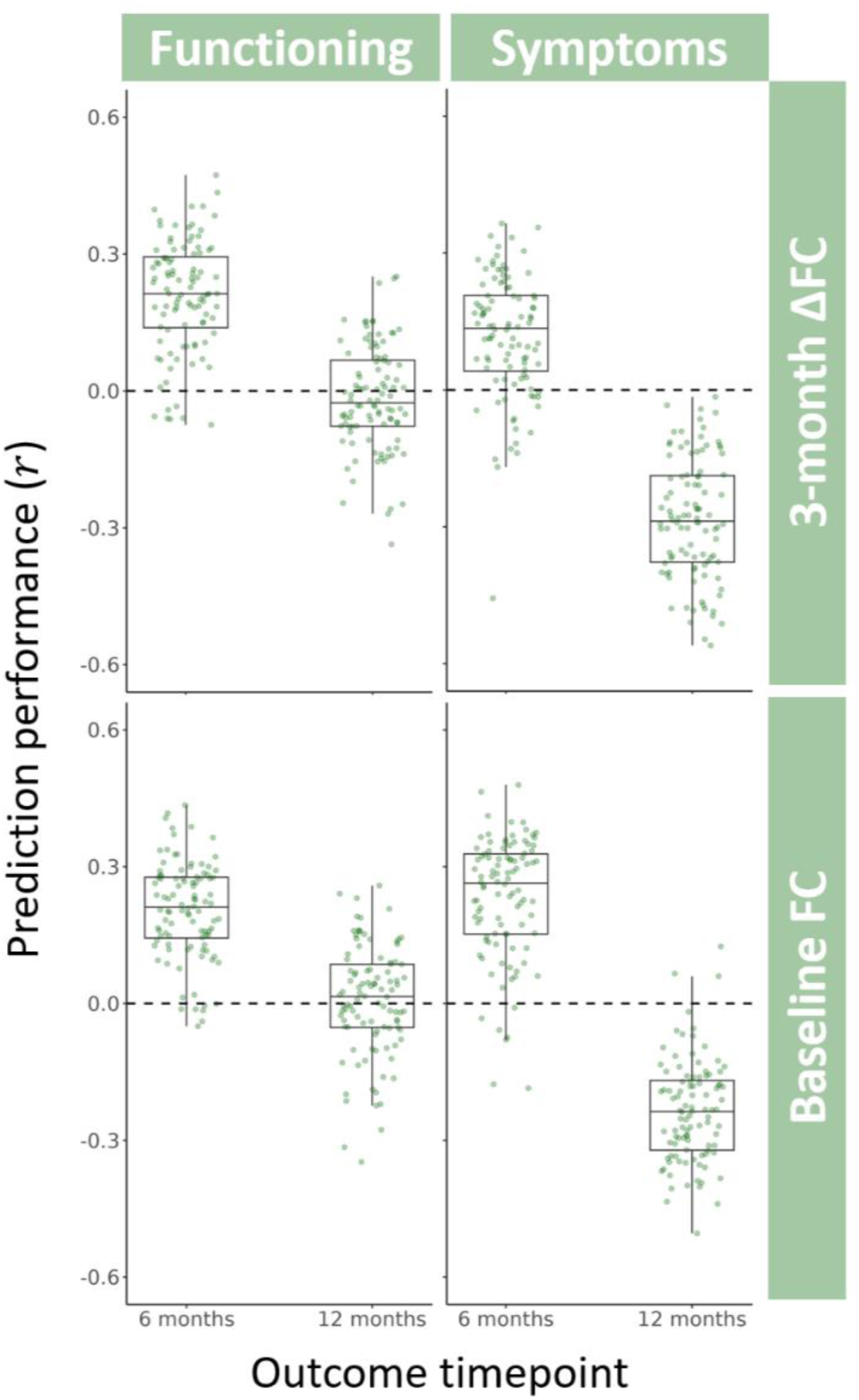
Performance of kernel ridge regression (KRR) for predicting patients’ 6- and 12-month changes in symptoms and functioning, using either baseline functional coupling (FC) or 3-month change in FC. Each data point shows the strength of Pearson’s correlation between predicted and observed clinical outcomes for a single split of 4 -fold cross-validation, with each of the eight models comprising 100 random splits.

**Table 2:**
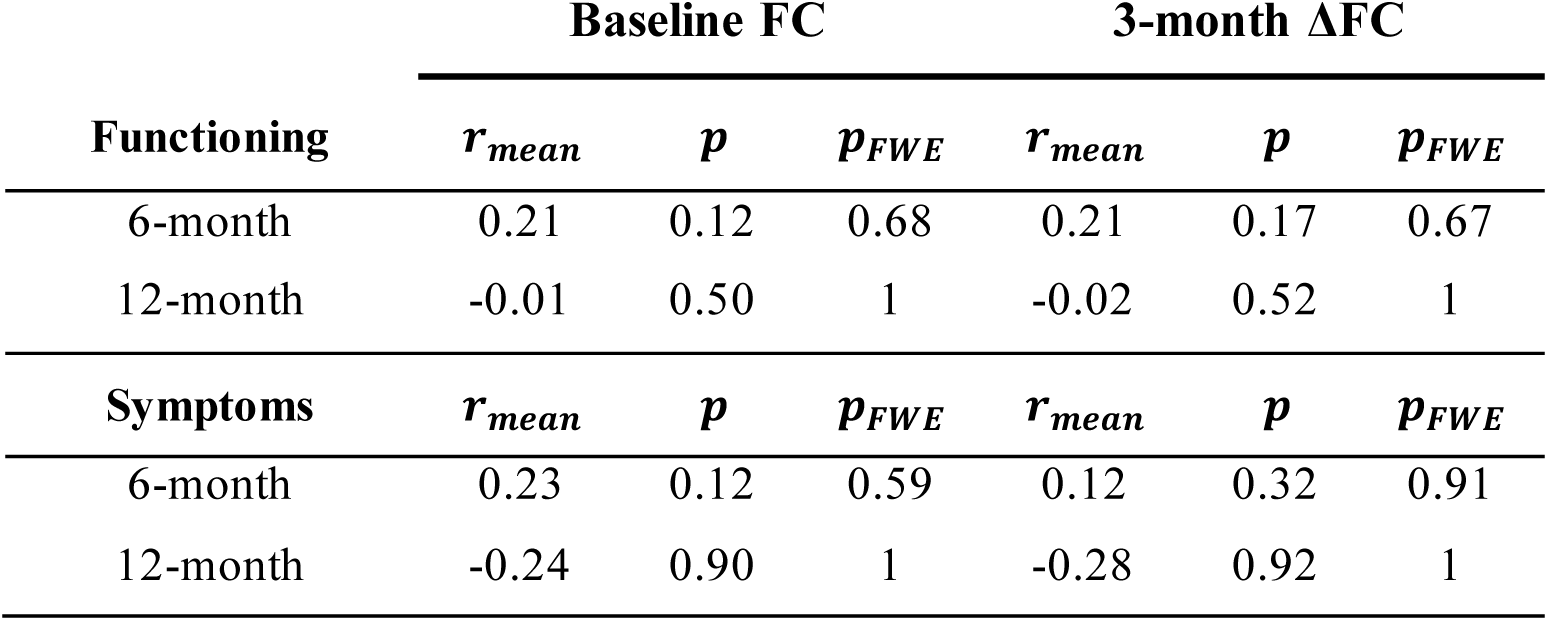
Prediction performance and significance for all eight kernel ridge regression (KRR) models.

#### Multilayer Meta-matching

The four multilayer meta-matching models that used baseline FC to predict outcomes also demonstrated limited performance (*r*_*mean*_ range of −0.16 to 0.24; Table 3 and Figure 4) and none achieved significance before or after FWE correction (all *p* ≥ 0.09; Table 3 and Figure S5).

**Figure 4:**
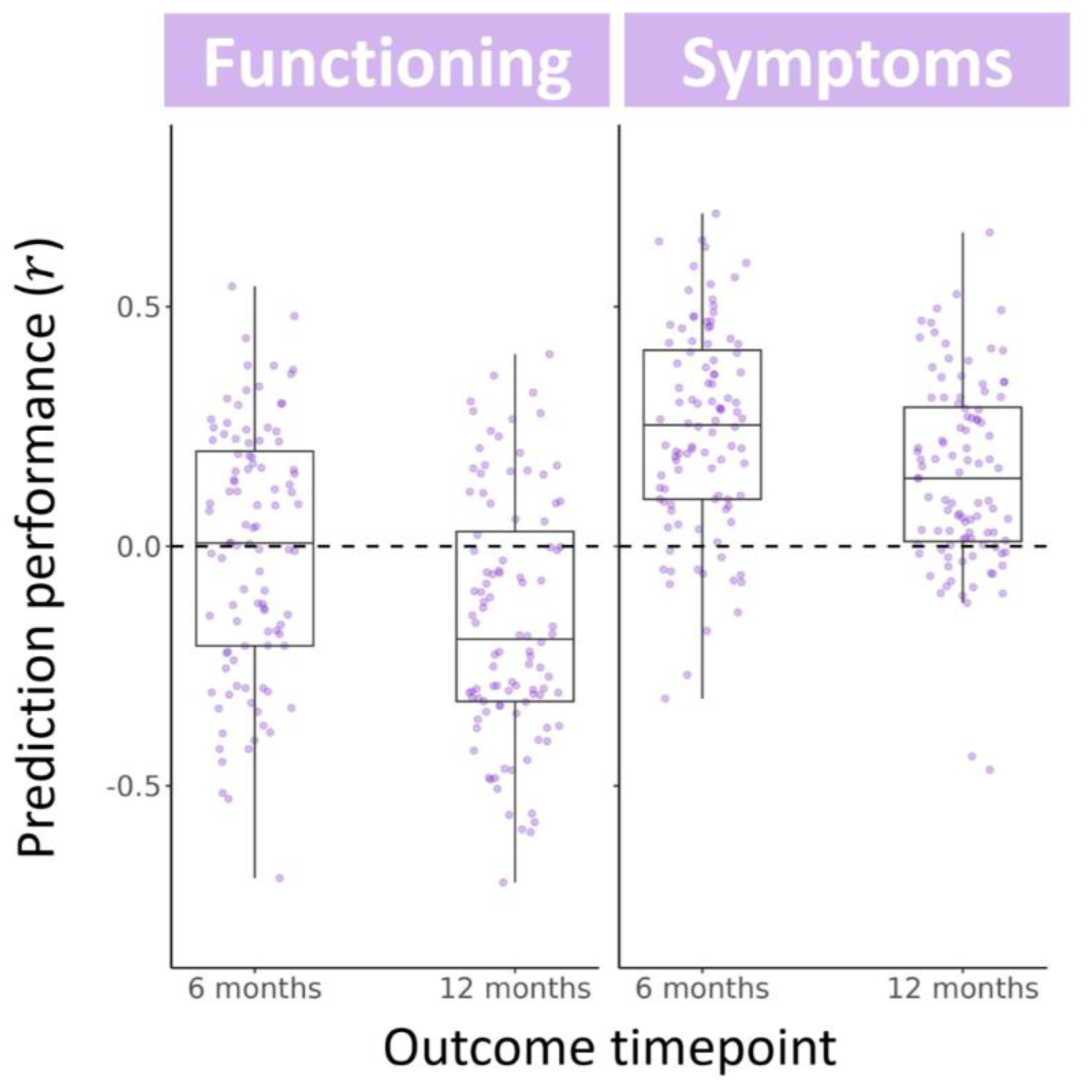
Performance of multilayer meta-matching for predicting patients’ 6- and 12-month changes in symptoms and functioning, using baseline functional coupling (FC). Each data point shows the strength of Pearson’s correlation between predicted and observed clinical outcomes for a single split of 4 -fold cross-validation, with each of the four models comprising 100 random splits.

**Table 3:** Prediction performance and significance for all four multilayer meta-matching models.

|  | Baseline FC |  |  |
| --- | --- | --- | --- |
| | $r_{mean}$ | $p$ | $p_{FWE}$ |
| **Functioning** |  |  |  |
| 6-month | -0.01 | 0.53 | 0.94 |
| 12-month | -0.16 | 0.83 | 1 |
| **Symptoms** |  |  |  |
| 6-month | 0.24 | 0.09 | 0.29 |
| 12-month | 0.15 | 0.21 | 0.58 |

## Discussion

Many fMRI studies have reported disrupted FC across the psychosis spectrum by comparing patients to healthy controls (18,20–28), yet it remains unclear whether FC holds potential as a prognostic biomarker that can predict treatment outcomes in FEP. We leveraged the long-term longitudinal measurement of symptoms and functioning from a clinical trial, along with gold-standard prediction algorithms, to evaluate whether resting-state FC could predict FEP patients’ outcomes at the end of the 6-month trial and after 12 months.

All the CPM, KRR, and multilayer meta-matching algorithms showed poor prediction performance (all *r*_*mean*_ < 0.3) for both outcome measures and timepoints, and no model achieved statistical significance. Our results contrast prior FEP prediction studies, where FC either predicted treatment response compared to non-response (defined via a priori symptom thresholds) with balanced accuracies exceeding 75% (39,40,42,43), or significantly predicted longitudinal changes in symptoms (38,41,42,44).Several differences between the prior and current work may explain this difference. First, our FEP patient sample included the full spectrumof psychoticdisorders, whereas most prior studies only included patients diagnosed with schizophrenia. Second, patients in our sample received psychosocial interventions alongside either placebo or antipsychotic tablets, whereas prior studies generally involved more rigid treatment protocols (e.g., all patients received the same antipsychotic dosing with no psychosocial interventions). Third, we considered whole-brain FC, whereas most prior studies defined a priori predictors localized to specific regions or networks. Predictions were consequently driven by different FC patterns across studies, including cerebello-thalamo-cortical (41), cortico-cortical (38,42), and hippocampal-cortical (39) circuits as well as FC seeded in the bilateral anterior cingulate cortex (44) and striatum (40).

When taken with our null findings, this body of work suggests that FC may only predict clinical outcomes within highly selected samples and contexts, and may not be generalizable to the heterogeneous samples and treatment settings encountered in real-world clinics (84,85). It is also possible that FC may only capture a patient’s capacity for clinical changes within the early phase of treatment, given that the only other study to predict outcomes beyond 16 weeks also reported the lowest performance (*r* = 0.21) amongst prior studies using continuous outcomes (41).

A separate study of this cohort used CPM with baseline measures of structural connectivity to significantly predict 12-month changes in functioning (*r*_*mean*_ = 0.44), significantly outperforming predictive models based on patients’ baseline clinical, cognitive, and demographic data (19). This result suggests that diffusion MRI-derived estimates of structural connectivity hold greater potential for a prognostic biomarker in FEP than resting-state FC, which has a lower test-retest reliability (86) and correlates with a range of in-scanner internal states (87–89) and self-reported experiences (90,91). However, prognostic trait-like FC patterns may be revealed by estimating FC during movie-watching (92) or across multiple task and rest conditions (93–96) [see Cao et al. (2023) for FEP outcome prediction based on such cross-paradigm FC (38)].

### Limitations

Although our sample size of *n* = 35-49 was similar to those featured in prior studies, it may be underpoweredfor robust predictive modelling (65–69). We included multilayer meta-matching to compensate for the small sample size, but this approach only improves prediction performance compared to single-sample methods when the phenotype of interest is strongly correlated withat least one phenotypefrom the large source datasets used in training (58,97). To our knowledge, no prior studies have used multilayer meta-matching for longitudinal prediction, and so patients’ clinical outcomes may not have been closely related to any of the 229 phenotypes in the five cross-sectional source datasets. The reliability and generalizability of our finding, that resting-state FC cannot predict FEP patients’ clinical outcomes, can therefore only be assessed by performing cross-validation on a larger sample (67,84,85). This remains challenging, however, due to the paucity of open-access FEP neuroimaging datasets with longitudinal outcomes.

The BPRS, which we used to measuresymptomseverity, demonstrates lower inter-rater reliability, internal consistency, and clinical predictive power than other inventories which require more burdensome administration (98,99). This may have weakened our statistical power for detecting FC-outcome associationsand explain the poorpredictionperformance for symptom changes compared to studies that used other scales (41–43). The construct validity of our symptom outcomes may have been limited by summing scores across the 18 items in the BPRS, as some psychometric literature suggests that sum-scores can obfuscate separatedimensionsof psychopathology in highly heterogenous disorders (100–102). In the present study, we chose our predicted outcomes to align with the preregistered clinical trial outcomes of STAGES and minimize FWE correction. To improve clinical relevance, future studies could instead predict changes within subscales demarcated to capture more homogenous symptom dimensions (e.g., depression, anxiety, psychotic symptoms) (103,104), or use factor analytic techniques to maximize effect sizes (101,105).

Heterogeneity in clinical outcomes amongst FEP patients can also be explained by non-neurobiological factors, including baseline symptoms and functioning (4,5,10,85,106–113), age (4,107,114,115), sex (4,10,110,112), schizotypal traits (10,107), inflammatory markers (116,117), education and employment status (109,110,115,118), substance use (109,112,119,120), recent life events (121), previous depressive episodes (110), social environment (110,122), and duration of untreated psychosis (4–6,106–108,113,123–127). If these factors do not covary with neuroimaging measures, their effects will impose an upper limit to the performance and clinical utility of brain-based prognostic biomarkers. It will be important for future work to evaluate how the combination of different biological, demographic, and clinical measures affects prediction of outcomes (128–130). Indeed, given the expense and complexity of fMRI, any promising FC-based prognostic biomarkers should demonstrate predictive capacity beyond that afforded by simpler measures (85,108,131,132).

## Conclusions

Our analysis, usingmultiple cross-validated prediction algorithms, indicatesthat brain-wideresting-state FC at baseline, or FC change over 3 months, does not significantly predict FEP patients’ changes in symptoms or functioning over a 6- to 12-month period after commencing treatment. When taken together with past work, this finding suggests that FC may only hold prognostic utility within narrow clinical and experimental contexts.

## Data Availability

All data produced in the present study are available upon reasonable request to the authors. Code to reproduce all main results figures is available at https://github.com/izachp/psychosis-FC-prediction.

## Acknowledgements

Janssen-Cilag partially supported the early years of this study with an unrestricted investigator-initiated grant and provided risperidone, paliperidone, and matched placebo for the first 30 participants. The study was then funded by an Australian National Health and Medical Research Project (NHMRC) grant (Grant No. 1064704 [to SMF, BO, BN, HPY, KA, MA-J, SH, SJW, PM, AF]). The funders had no role in study design, data collection, data analysis, data interpretation, writing, approval, or submission of this manuscript. No patients or members of the public were involved in the design, conduct, reporting, interpretation, or dissemination of the study.

This study has been supported by a large number of clinical staff at Orygen Youth Health (Craig Macneil, Kingsley Crisp, Dylan Alexander, Tina Proffitt, Rachel Tindall, Jennifer Hall, Lisa Rumney, Franco Scalzo, Melissa Pane, Linda Kader, Frank Hughes, Clare Shelton, Ryan Kaplan, David Hallford, Bridget Moller, and Rick Fraser) and research assistants (Daniela Cagliarini, Suzanne Wiltink, Janine Ward, Sumudu Mallawaarachichi, Lara Baldwin, and Jessica Graham). The trial took place at the Early Psychosis Prevention and Intervention Centre, which is part of Orygen Youth Health, Melbourne, Australia.

SC is supported by a Graduate Education Fund Scholarship from the American Australian Association. SF, MA-J, and AF reported receiving grants from the NHMRC and Australian Research Council during the conduct of the study. CP reported receiving grants from the Australian NHMRC and from the Lundbeck Foundation and personal fees from Lundbeck Australia Pty Ltd. Advisory Board for talks presented at educational meetings organized by Lundbeck. PM reported receiving grants from the Australian NHMRC, the Colonial Foundation, the National Alliance for Research on Schizophrenia and Depression, the Stanley Foundation, the National Institutes of Health, Wellcome Trust, the Australian and Victorian governments, and Janssen-Cilag (unrestricted investigator-initiated grant) during the conduct of the study; past unrestricted grant funding from Janssen-Cilag, AstraZeneca, Eli Lilly, Novartis, and Pfizer; and honoraria for consultancy and teaching from Janssen-Cilag, Eli Lilly, Pfizer, AstraZeneca, Roche, Bristol Myers Squibb, and Lundbeck. BN was supported by an NHMRC Senior Research Fellowship (Fellowship No. 1137687) and VLC was supported by an NHMRCEL2 Fellowship (Fellowship No. 1177370). BN, KA, and VLC were supported by a University of Melbourne Dame Kate Campbell Fellowship.

This work was supported by the MASSIVE HPC facility (www.massive.org.au) (133). All code used for statistical analyses and generating main results figures can be found at https://github.com/izachp/psychosis-FC-prediction.

## Disclosures

Dr Francey reported receiving grants from the Australian National Health & Medical Research Council (NHMRC) during the conduct of the study. Dr Allott reported receiving grants from the Australian National Health and Medical Research Council during the conduct of the study. Dr Pantelis reported receiving grants from the Australian NHMRC and from the Lundbeck Foundation and personal fees from Lundbeck Australia Pty Ltd Advisory Board for talks presented at educational meetings organized by Lundbeck. Dr McGorry reported receiving grants from the Australian NHMRC, the Colonial Foundation, the National Alliance for Research on Schizophrenia and Depression, the Stanley Foundation, the National Institutes of Health, Wellcome Trust, the Australian and Victorian governments, and Janssen-Cilag (unrestricted investigator-initiated grant) during the conduct of the study; past unrestricted grant funding from Janssen-Cilag, AstraZeneca, Eli Lilly, Novartis, and Pfizer; honoraria for consultancy and teaching from Janssen-Cilag, Eli Lilly, Pfizer, AstraZeneca, Roche, Bristol Myers Squibb, and Lundbeck. Dr Fornito received grants from the National Health and Medical Research Council, Australian Research Council, and Sylvia and Charles Viertel Foundation during this project. No other disclosures were reported.

## Notes

### Clinical Trial

ACTRN12607000608460

### Author Declarations

The current study uses data from a larger trial registered with the Australian New Zealand Clinical Trials Registry in November 2007 (ACTRN12607000608460) and received ethics approval from the Melbourne Health Human Research and Ethics committee.

### Summary of Updates

Improved figure resolutions and PDF formatting.

