## Supplemental Information for "Functional Coupling and Longitudinal Outcome Prediction in First-Episode Psychosis"

### Supplement


Supplementary Information – STAGES clinical trial details

Supplementary Information – MRI acquisition, quality control, and preprocessing details

Supplementary Information – Further information on kernel ridge regression

Supplementary Table 1 – Sample characteristics for included patients

Supplementary Figure 1 – Quality control metrics throughout fMRI preprocessing

Supplementary Figure 2 – Clinical outcomes for included patients

Supplementary Figure 3 – Null distribution for connectome-based predictive modelling

Supplementary Figure 4 – Null distribution for kernel ridge regression

Supplementary Figure 5 – Null distribution for multilayer meta-matching

Supplementary Figure 6 – Performance of connectome-based predictive modelling with feature selection at $p < 0.05$

Supplementary Figure 7 – Null distribution for connectome-based predictive modelling with feature selection at $p < 0.05$

Supplementary Table 2 – Prediction statistics for connectome-based predictive modelling with feature selection at $p < 0.05$

Supplementary Figure 8 – Performance of connectome-based predictive modelling with feature selection at $p < 0.001$

Supplementary Figure 9 – Null distribution for connectome-based predictive modelling with feature selection at $p < 0.001$

Supplementary Table 3 – Prediction statistics for connectome-based predictive modelling with feature selection at $p < 0.001$

**STAGES clinical trial details**

To maximize safety and comply with standards set by the Melbourne Health Human Research and Ethics Committee, potential participants were subject to the following exclusion criteria: (i) inability to provide informed consent; (ii) risk to self or others (score of 5 or greater on the BPRS-4 Suicidality and Hostility subscales); (iii) duration of untreated psychosis greater than 6 months; (iv) pregnancy; (v) non-negligible prior use of antipsychotics outside the trial (7 days or 1750 mg chlorpromazine equivalent) or present use of mood stabilizers; and (vi) unstable accommodation or support. Study discontinuation criteria also included these items, along with (vii) substantial exacerbation of positive symptoms (2-point increase on the BPRS-4 subscale for Conceptual Disorganization, Hallucinations, Unusual Thought Content, or Suspiciousness, maintained for 1 week and not due to substance use); (viii) substantial and sustained drop in overall functioning (20-point decrease on the SOFAS from baseline, maintained for 1 month); (ix) request by the participant for antipsychotic medication; and (x) minimal recovery after 3 months (>4 on the BPRS-4 Hallucinations, Suspiciousness, and Unusual Thought Content subscales, or >3 on Conceptual Disorganization).

Participants were allocated to antipsychotic and placebo arms of the trial in a 1:1 ratio, using a randomization design that stratified for sex and the duration of untreated psychosis (0-30, 31-90, >90 days). Participants, clinicians, and research staff remained blinded to this allocation throughout the trial. Participants in the antipsychotic arm were administered either 1 mg risperidone or 3 mg paliperidone, depending on recruitment period. Multiple compounds were used due to changing availabilities of matched placebo tablets, with these dosages converted using olanzapine equivalent units. All participants received cognitive behavioral case management, comprising manualized cognitive behavioral therapy and psychoeducation, aimed at preventing relapse and enhancing coping strategies for positive and negative symptoms (1). Additional support via group programs and advocacy was provided by case managers. Clinicians assessed participants' BPRS and SOFAS scores at each session. After the 6-month treatment period, all participants were offered open-label treatment at the same clinic. Participants who discontinued the trial were offered to remain in cognitive behavioral case management and receive antipsychotic medication as deemed appropriate by the treating team. The full study protocol can be found in O'Donoghue et al. (2).

**MRI acquisition, quality control, and preprocessing details**

Participants were instructed to maintain wakefulness and remain still while lying in the scanner with their eyes open. For each anatomical scan, interleaved acquisition was used to capture 176 slices under the following parameters: repetition time = 2300ms; echo time = 2.98ms; flip angle of 9°; FOV of 256mm; voxel size of 1.1 x 1.1. x 1.2 mm. For each functional scan, interleaved acquisition was used to capture 37 slices per volume, for 234 volumes, under the following parameters: repetition time = 2000ms; echo time = 32ms; flip angle = 90°; field of view = 210mm; slice thickness of 3.5 mm, and 3.3 x 3.3 x 3.55 mm voxels.

A total of 98 scans were first evaluated by the quality control software MRI-QC, which automatically estimates 56 no-reference image quality metrics for T1-weighted scans and 31 metrics for functional scans. By providing both individual- and group-level visualizations of these metrics, MRI-QC allows the identification of any outlier scans with issues such as motion artefacts, signal spillover, coil artefacts, poor contrast, and signal dropout (3). We excluded three problematic scans, then preprocessed remaining scans using the standard automated pipeline in fMRIPrep v1.4.1 (4) as follows.

First, anatomical images were corrected for signal intensity non-uniformity via the N4BiasFieldCorrection algorithm, to improve tissue contrast across the brain (5). All 176 slices per scan were then spatially aligned to each other in a common orientation using the Advanced Normalization Tools (ANTs) package (6). Images were also skull-stripped with ANTs, prior to tissue segmentation using FMRIB's Automated Segmentation Tool (7). FreeSurfer (8) was used to reconstruct pial, grey matter, and white matter surfaces from these tissue maps. Anatomical volumes were then transformed to the standard MNI152NLin6Asym space (9) using ANTs. Separately, all 37 slices per functional volume were spatially aligned to each other, then all 234 volumes per scan were spatially aligned to each other. This corrects for in-scanner head motion, with brain realignment parameters estimated in 6 time series (translation and rotation in 3 dimensions each) by the MCFLIRT function in the FMRIB software library (10). To adjust for the asynchronous acquisition of slices, the AFNI package performed slice-time correction (11), which temporally shifts the blood oxygen level-dependent (BOLD) signal time series of each slice to adjust for the asynchronous acquisition of slices. This step aligns with recommendations by Sladky et al. (2011), who found robust increases in statistical power for repetition times of 2 seconds or longer (12). To adjust for inhomogeneities in the applied magnetic field, which are known to stretch, compress, and shift image voxels (13), ANTs performed susceptibility distortion estimation and correction. FreeSurfer then aligned all functional volumes to the anatomical volumes.

In-scanner head motion has been shown to distort blood oxygen level dependent (BOLD) signal time series, with spatially heterogenous effects leading to spurious increases and decreases in estimates of FC (14–16). We used the method from Jenkinson et al. (2002) to calculate each fMRI scan's time series of framewise displacement (FD) (10), which quantifies the total amount of head movement between each of the 234 volume acquisitions. We then classified and excluded one high-motion scan according to the stringent criteria in Parkes et al. (2018) (17), which advises to exclude a scan if it meets any of the following: (i) $\text{mean FD} > 0.25\ \text{mm}$; (ii) $20\%\ \text{of FD} > 0.2\ \text{mm}$; and (iii) any single $\text{FD} > 5\ \text{mm}$. The 94 remaining scans displayed low head motion ($\text{mean FD} = 0.049\ \text{mm}, \text{SD} = 0.020\ \text{mm}$) compared to large resting-state fMRI datasets of the general population (17).

Since fMRI has an inherently low signal-to-noise ratio, FC-based analyses typically include denoising beyond the minimal steps in fMRIPrep (17). We chose a denoising pipeline that has been shown to minimize three quality control benchmarks (17): (i) FD-FC correlations, which quantify how head motion impacts each FC estimate across all scans; (ii) FD-FC distance dependence, which quantifies how FD-FC correlations change with the coupling distance that separates each pair of regions; and (iii) temporal degrees of freedom lost by denoising, which confers reduced statistical power for analyses.

In each scan, we first applied linear detrending to the BOLD signal time series of every voxel. Scans were then denoised with independent component analysis for the automatic removal of motion artefacts (ICA-AROMA). ICA-AROMA identifies motion-related spatial components of the fMRI data based on a priori thresholds for spatial features (overlap with cerebrospinal fluid, overlap with pial surface) and a linear discriminant analysis for temporal features (correlation with head motion, high-frequency content) (18). In line with recommendations (18,19), smoothed fMRI data (kernel of 6 mm full-width at half-maximum) were input to ICA-AROMA. To mitigate the impacts of head motion on FC estimates, the time series of these motion-related components were then non-aggressively regressed out from every voxel in the non-smoothed detrended scan. Here, non-aggressive regression refers to performing regression while preserving any temporal variance shared between regressors (i.e., motion-related components) and signal (i.e., grey matter) time series, and was used in the original ICA-AROMA paper (18). To reduce the impact of any non-neural fluctuations (e.g., cardiac, respiratory) on FC estimation, we also regressed out mean signals of the white matter and cerebrospinal fluid tissues (20). These signals were calculated using conservative tissue masks, derived by repeatedly eroding the tissue probability maps from fMRIPrep (5 erosion cycles for white matter, 2 for cerebrospinal fluid).

One common yet controversial denoising step in resting-state fMRI preprocessing is global signal regression, which removes the mean whole-brain signal from every voxel (21). Since it centers each scan's FC distribution around zero, global signal regression introduces potentially spurious anti-correlations. Although the global signal partially reflects non-neural physiology and head motion (17,20,22), it also has behaviorally-relevant neuronal contributions (23–25). Importantly, global signal regression tends to improve behavioral prediction (26), justifying its inclusion in our study. Since the global signal is strongly correlated with the mean grey matter signal (21), we used grey matter regression as a substitute that may avoid reintroducing noise by regressing out white matter and cerebrospinal fluid signals twice (27). Grey matter tissue masks were derived by thresholding the corresponding fMRIPrep probability maps at 70%. As per recommendations (14,28), we also regressed out the temporal derivative (calculated as backward differences), the square, and the square of the temporal derivative for each of the white matter, cerebrospinal fluid, and grey matter time series. To avoid reintroducing noise, we calculated all tissue signals after ICA-AROMA denoising and performed regressions simultaneously (29). To further isolate neural signal, we applied a band-pass filter of $f = 0.008$-$0.08\ Hz$ to the BOLD signal time series.

For the CPM and KRR models, we parcellated each scan's grey matter using the 300-region cortical atlas from Schaefer et al. (2018) (30) and the 32-region subcortical atlas from Tian et al. (2020) (31). To adjust for individual differences in cortical anatomy, the Schaefer atlas was mapped onto each participant's grey matter using manually-corrected tissue surfaces from FreeSurfer. Regional BOLD signal time series were calculated as a mean across all voxels in each of the 332 regions, weighting each voxel's contribution by its fMRIPrep-estimated probability of being grey matter. To identify any regions with poor signal, we calculated each region's mean BOLD signal across all timepoints and scans. Using the method in Brown et al. (2019) (32), we excluded the 4 globi pallidi regions as their signals fell below the largest gap (or 'elbow') in the distribution of all 332 regions' signals. We did not remove any regions from the 419-region parcellation used for multilayer meta-matching, as this method requires complete data (33). Each scan's FC matrix was constructed by taking Pearson's correlation between the BOLD time series of each pair of regions.

FD-FC correlations were computed for all FC estimates in the 328-region parcellation after each denoising step, shown as box plots in Figure S1a. Despite the minimal head motion, moderate FD-FC correlations were present after fMRIPrep ($\text{mean}\ r = 0.11$, $\text{SD} = 0.12$), and were exacerbated by linear detrending ($\text{mean}\ r = 0.16$, $\text{SD} = 0.12$). Contrary to its intended purpose, ICA-AROMA had little impact on FD-FC correlations ($\text{mean}\ r = 0.17, \text{SD} = 0.11$), which were instead reduced by tissue-based regressions (8Phys-4GMR; $\text{mean}\ r = 0.014$, $\text{SD} = 0.13$) and band-pass frequency filtering ($\text{mean}\ r = 0.0061$, $\text{SD} = 0.12$). Moderate FD-FC distance dependence was present after fMRIPrep ($\rho = 0.19$; Figure S1b). Most subsequent denoising steps did not mitigate FD-FC distance dependence ($\rho = 0.21 - 0.22$), except band-pass filtering which slightly reduced the association ($\rho = 0.17$).

**Further information on kernel ridge regression**

Each kernel ridge regression model randomly divided patients into four folds, three of which formed the training set of $N$ individuals. For each individual $s$ in the testing fold, their clinical outcome $y_s$ was predicted as a weighted mean of the outcomes observed in the training set (contained in the $\boldsymbol{y}^{train}$ vector):

$$y_s = \boldsymbol{K}_s(\boldsymbol{K} + \lambda \boldsymbol{I})^{-1}\boldsymbol{y}^{\text{train}} \tag{1}$$

Here, $\boldsymbol{I}$ denotes the $N \times N$ identity matrix and $\boldsymbol{K}$ denotes the $N \times N$ matrix containing strengths of Pearson's correlation between every pair of vectorized upper-triangle FC matrices in the training set. That is, the $(i,j)^{th}$ element of $\boldsymbol{K}$ is given by:

$$(\boldsymbol{K})_{ij} = \text{corr}\left(FC_i^{\text{train}}, FC_j^{\text{train}}\right) \tag{2}$$

$\boldsymbol{K}_s$ similarly denotes the $1 \times N$ vector containing strengths of Pearson's correlation between the vectorized upper-triangle $FC$ matrix of individual $s$ and those in the training set:

$$(\boldsymbol{K}_s)_i = \text{corr}\left(FC_s, FC_i^{\text{train}}\right) \tag{3}$$

A range of values for the $l_2$-regularization hyperparameter $\lambda$ were tested via an inner loop of 4-fold cross-validation within the training set. The value of $\lambda$ which best minimised a cost function across individuals was then input in Equation 1 to predict outcomes in the testing set. More information is available at:

https://github.com/ThomasYeoLab/CBIG/tree/master/utilities/matlab/predictive_models/KernelRidgeRegression

**Table S1:** Baseline sample characteristics for STAGES patients included in the present study.

| | **First-episode psychosis patients ($n = 55$)** |
|---|---|
| Baseline age, years (SD) | 19.24 (2.86) |
| Females, N (%) | 28 (50.9%) |
| Left handedness, N (%) | 3 (5.5%) |
| Education, years (SD) | 12.22 (2.15) |
| Diagnosis, N | |
| Major depression with psychosis | 11 |
| Schizophreniform disorder | 8 |
| Psychotic disorder not otherwise specified | 14 |
| Substance-induced psychotic disorder | 6 |
| Delusional disorder | 5 |
| Schizophrenia | 10 |
| Missing diagnosis | 1 |
| Baseline BPRS total, mean (SD) | 56.76 (9.62) |
| Baseline SOFAS, mean (SD) | 52.49 (12.31) |
| Baseline SANS, mean (SD) | 34.07 (17.25) |
| Baseline HAM-D, mean (SD) | 18.87 (6.04) |
| Baseline HAM-A, mean (SD) | 20.73 (6.57) |
| Baseline QLS, mean (SD) | 69.42 (22.24) |

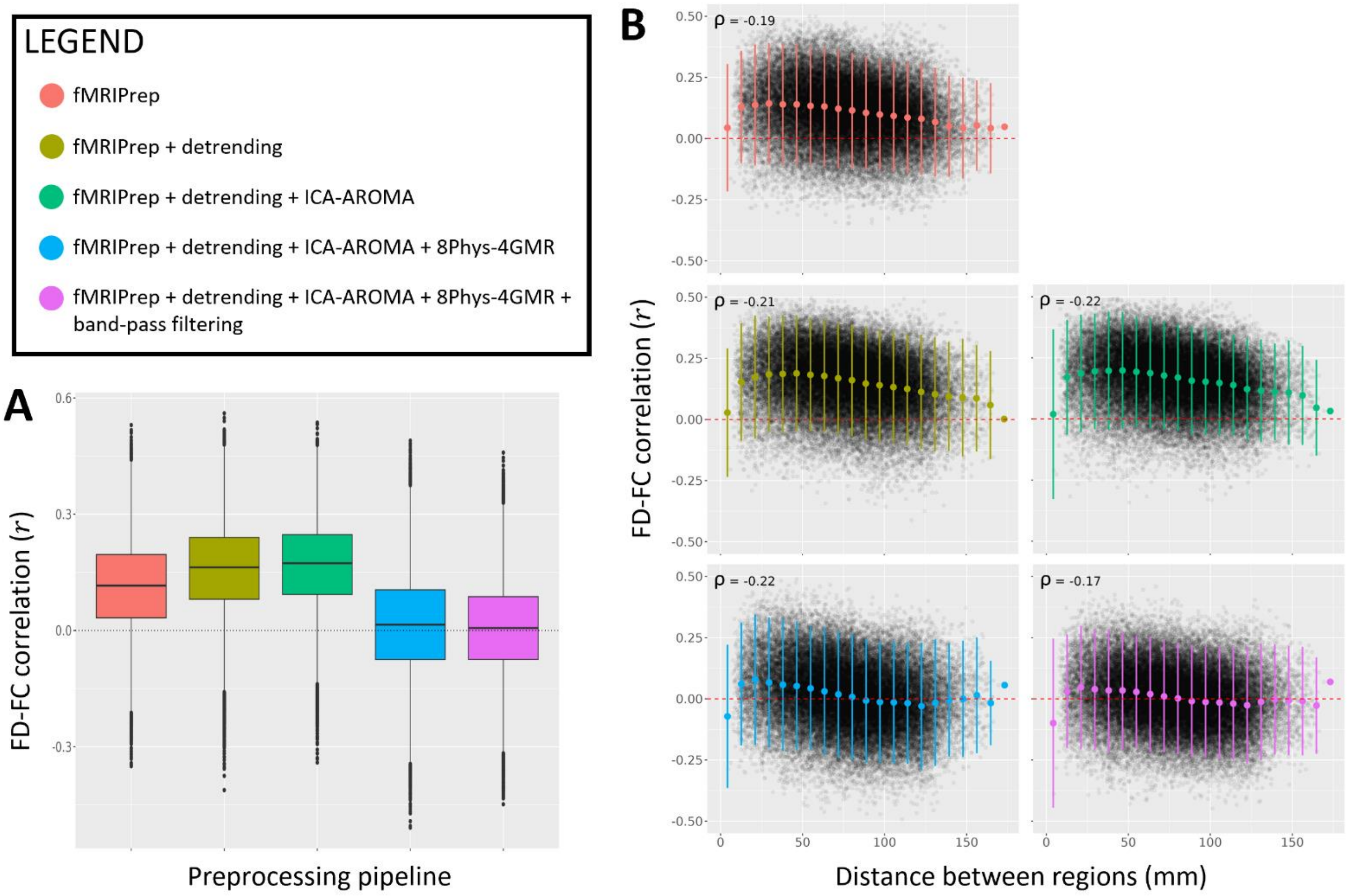


**Figure S1:** Quality control metrics quantifying the impact of in-scanner head motion on FC estimates, assessed throughout fMRI denoising. **(a)** FD-FC correlation distributions, computed after each denoising step outlined in the legend. Each data point represents the FD-FC correlation for a single FC estimate, across all scans. **(b)** FD-FC correlations plotted against the coupling distance that separates each pair of regions. Each plot was computed after a different denoising step, with mean and standard deviation bars colored according to the legend. The FD-FC distance dependence is indicated by Spearman's rho on each plot. '8Phys-4GMR' refers to the tissue-based regressions.

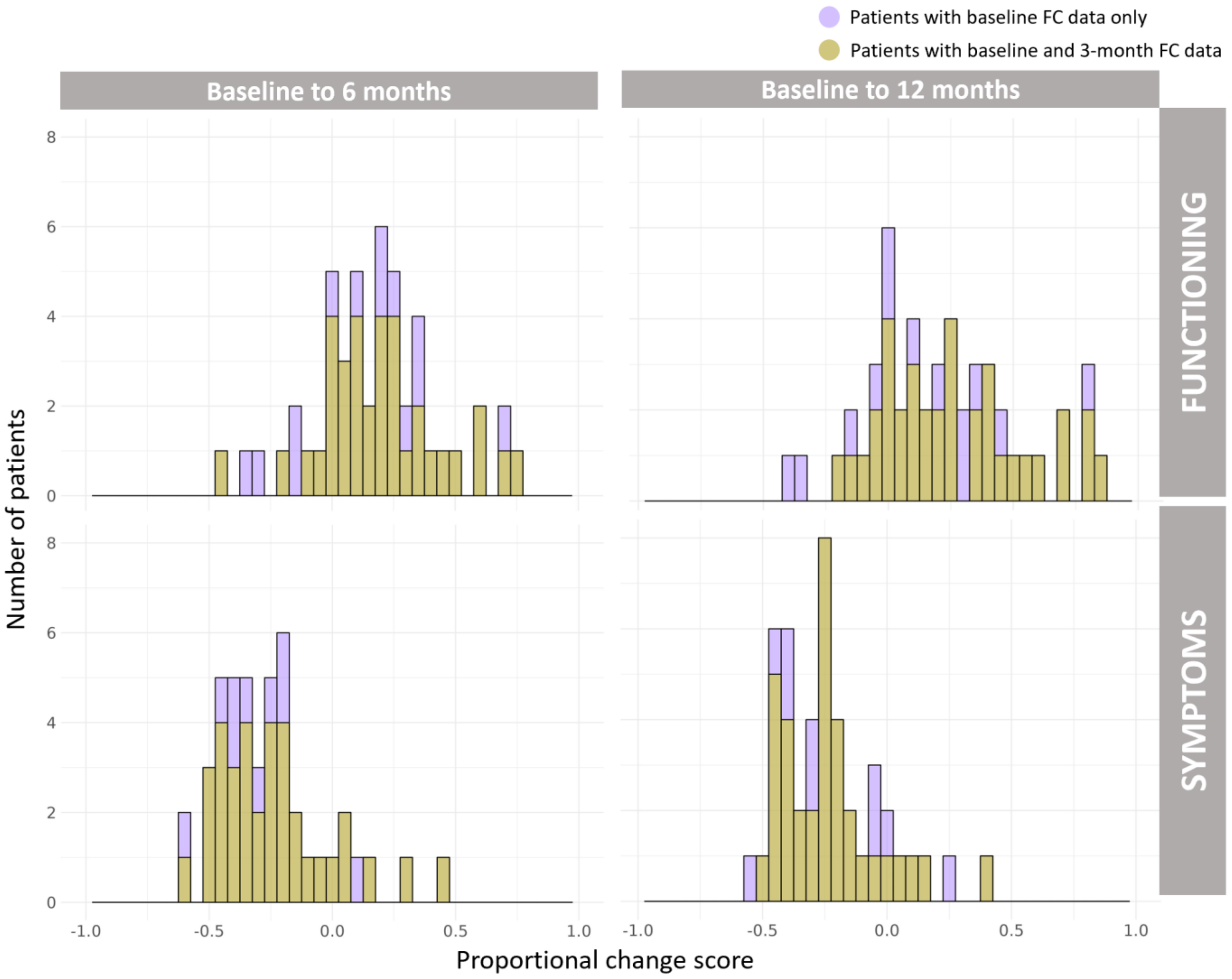


**Figure S2:** Clinical outcomes for patients included in the present study, defined as proportional changes in total SOFAS and BPRS scores after 6 and 12 months.

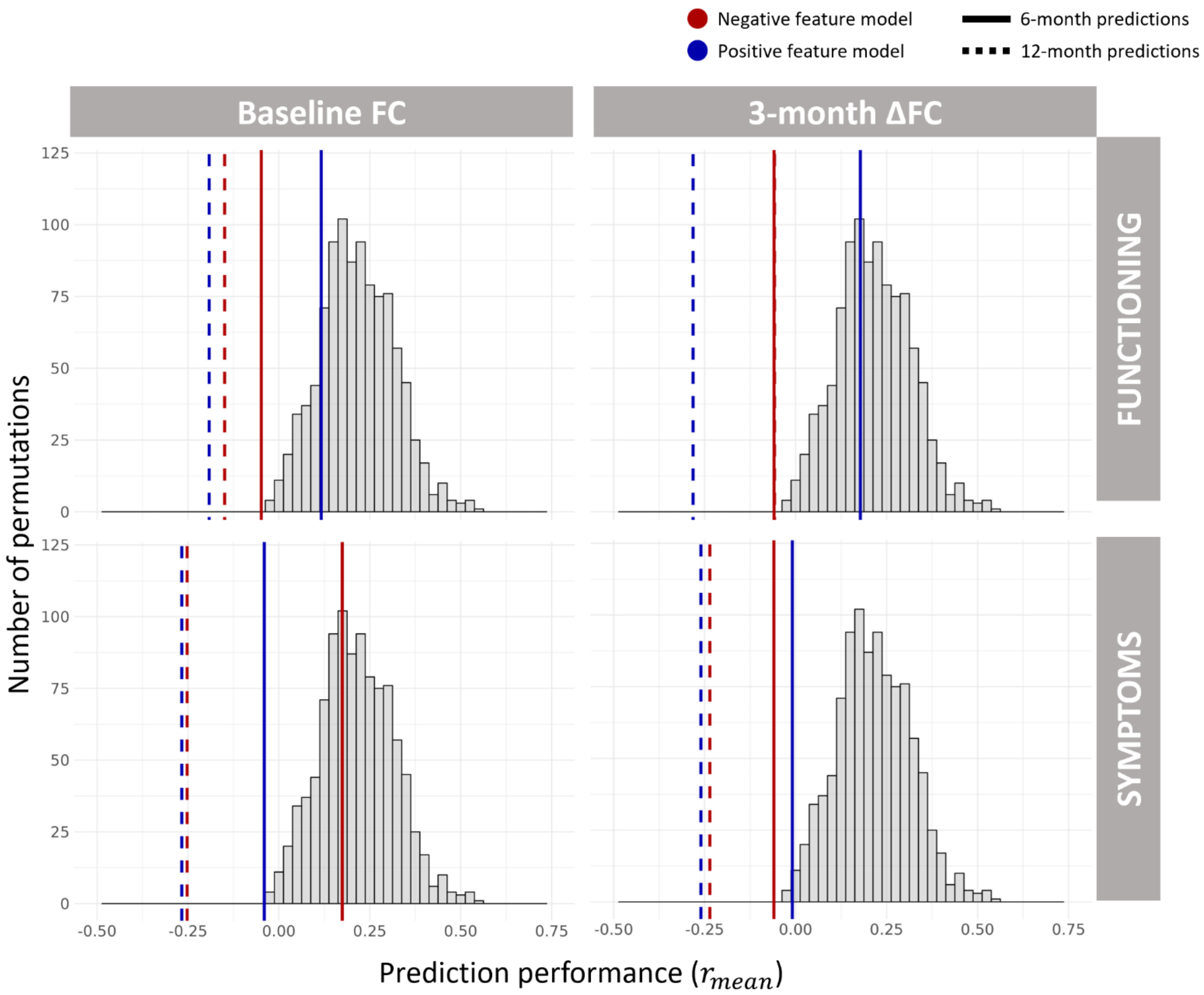


**Figure S3:** Prediction performance ($r_{mean}$) for connectome-based predictive modelling (CPM), shown in blue and red, superimposed against a family-wise error (FWE)-corrected empirical null distribution derived by randomly permuting clinical outcomes amongst patients. For each of 1000 permutations, 100 splits of CPM were run to calculate a single null $r_{mean}$.

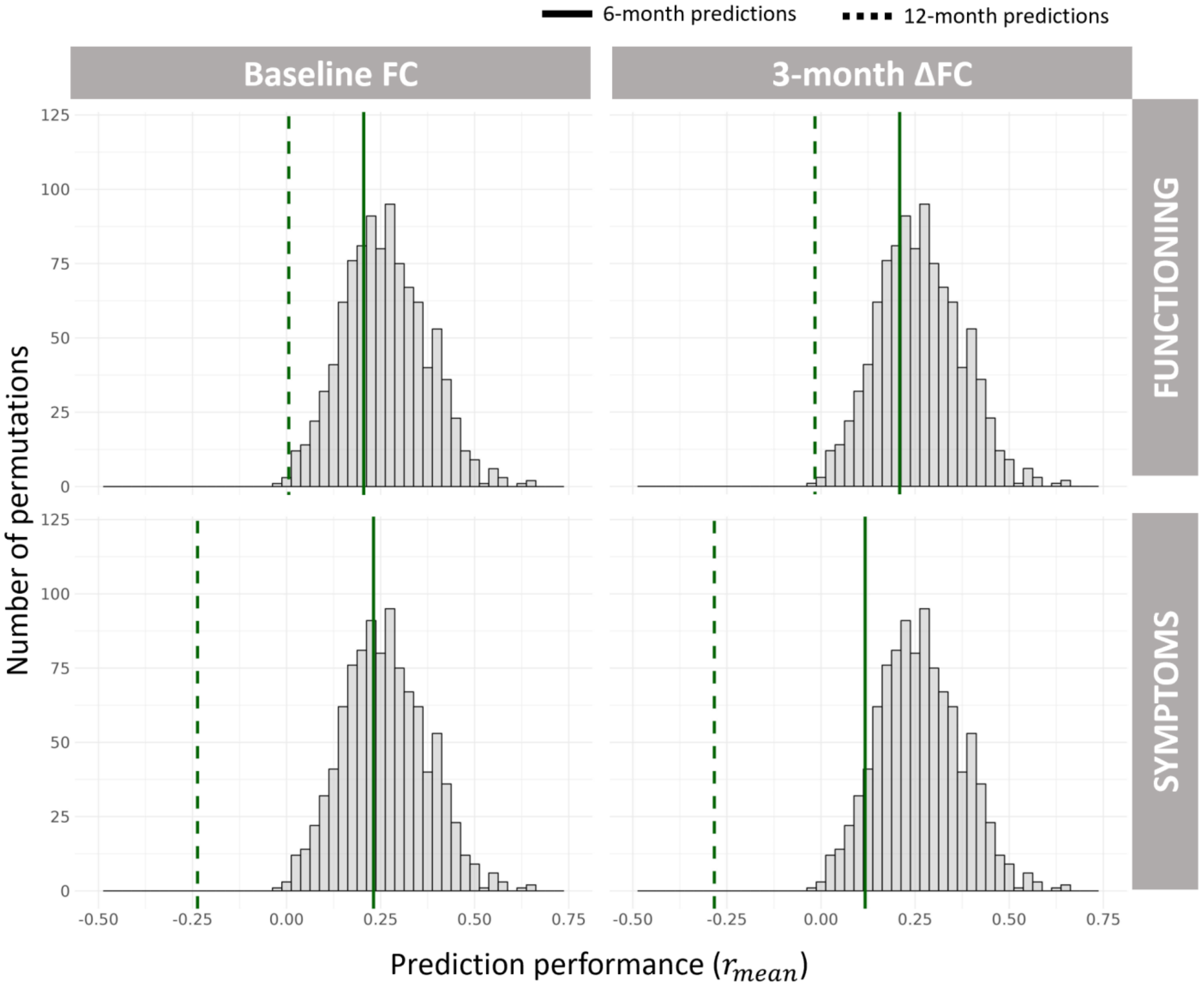


**Figure S4:** Prediction performance ($r_{mean}$) for kernel ridge regression (KRR), shown in green, superimposed against a family-wise error (FWE)-corrected empirical null distribution derived by randomly permuting clinical outcomes amongst patients. For each of 1000 permutations, 50 splits of KRR were run to calculate a single null $r_{mean}$.

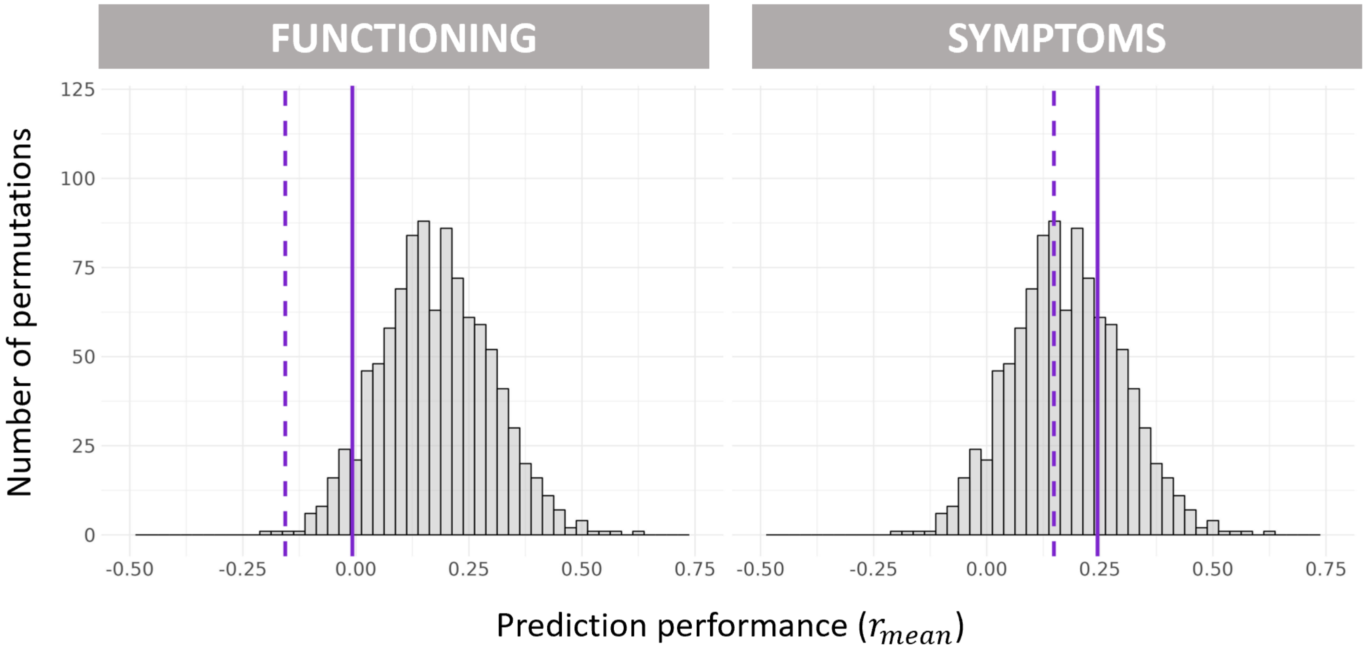


**Figure S5:** Prediction performance ($r_{mean}$) for multilayer meta-matching, shown in purple, superimposed against a family-wise error (FWE)-corrected empirical null distribution derived by randomly permuting clinical outcomes amongst patients. For each of 1000 permutations, 20 splits of multilayer meta-matching were run to calculate a single null $r_{mean}$.

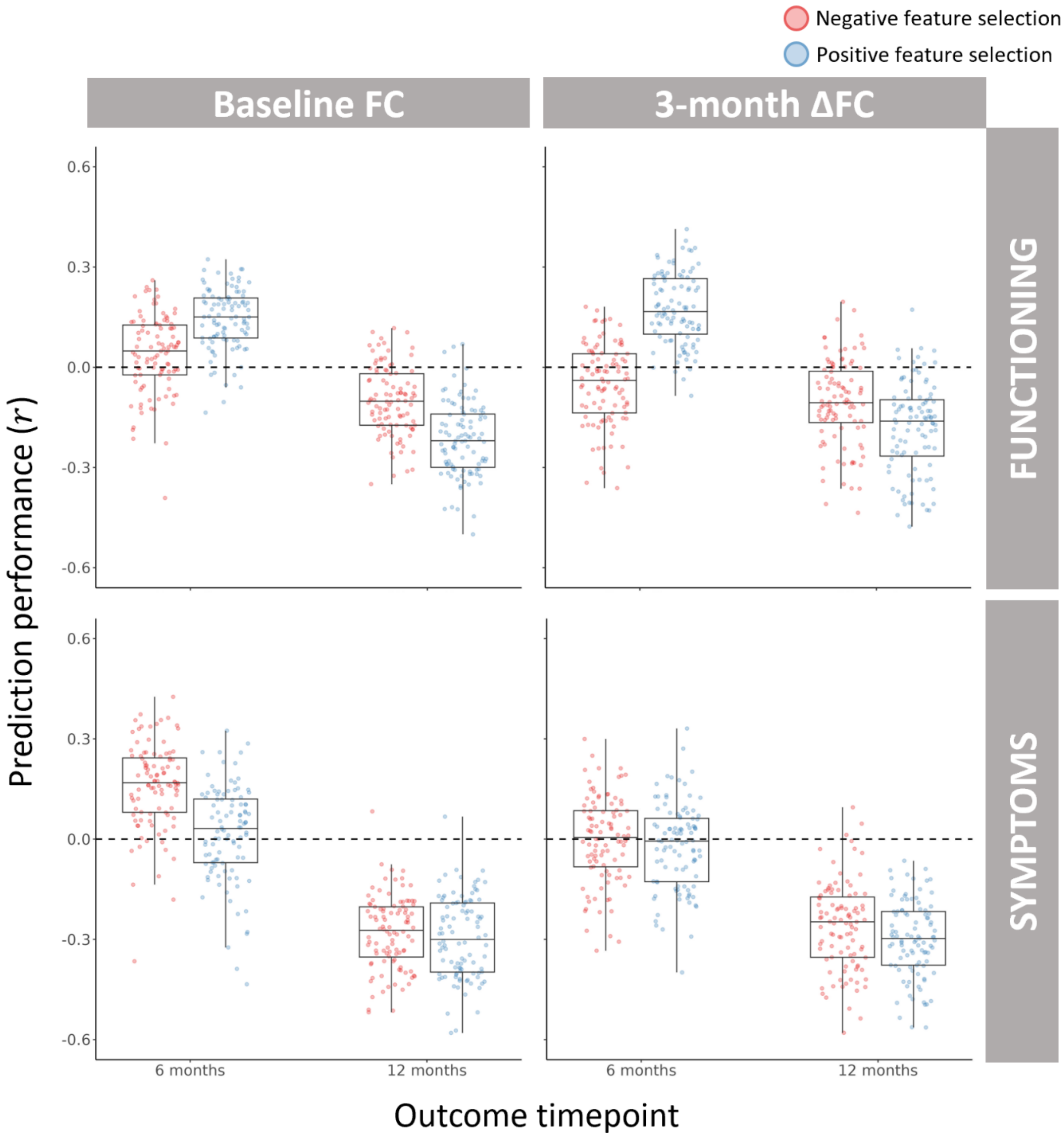


**Figure S6:** Performance of connectome-based predictive modelling (CPM) for predicting patients' clinical outcomes, using an alternate feature selection threshold of $p < 0.05$. Each data point shows the strength of Pearson's correlation between predicted and observed clinical outcomes for a single split of 4-fold cross-validation, with each of the 16 models comprising 100 random splits.

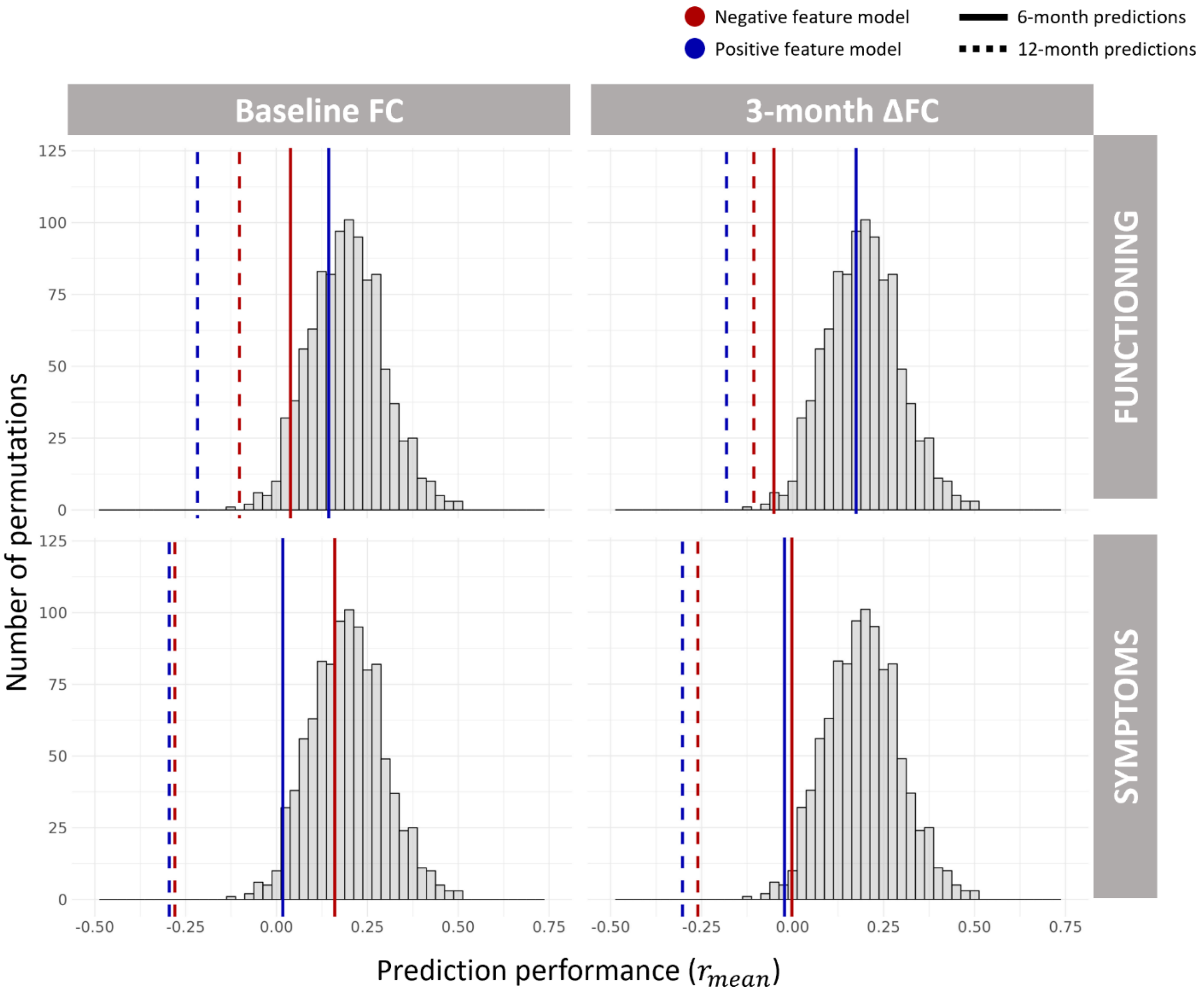


**Figure S7:** Prediction performance ($r_{mean}$ shown in red and blue) for all 16 connectome-based predictive modelling (CPM) models, with an alternate feature selection threshold of $p < 0.05$. Model performances are superimposed against a family-wise error (FWE)-corrected empirical null distribution derived by randomly permuting clinical outcomes amongst patients. For each of 1000 permutations, 100 splits of CPM were run to calculate a single null $r_{mean}$. FWE correction was applied across the 16 models shown.

**Table S2:** Prediction performance and significance for all 16 connectome-based predictive modelling (CPM) models, with an alternate feature selection threshold of $p < 0.05$. pos, positive feature model; neg, negative feature model.

| | | **Baseline FC** | | | **3-month ΔFC** | | |
|---|---|---|---|---|---|---|---|
| **SOFAS predictions** | | $r_{mean}$ | $p$ | $p_{FWE}$ | $r_{mean}$ | $p$ | $p_{FWE}$ |
| *6-month* | *pos* | 0.14 | 0.08 | 0.68 | 0.17 | 0.06 | 0.57 |
| | *neg* | 0.04 | 0.26 | 0.94 | -0.05 | 0.36 | 0.99 |
| *12-month* | *pos* | -0.22 | 0.79 | 1 | -0.18 | 0.64 | 1 |
| | *neg* | -0.10 | 0.52 | 1 | -0.11 | 0.48 | 1 |
| **BPRS predictions** | | $r_{mean}$ | $p$ | $p_{FWE}$ | $r_{mean}$ | $p$ | $p_{FWE}$ |
| *6-month* | *pos* | 0.02 | 0.27 | 0.97 | -0.02 | 0.31 | 0.99 |
| | *neg* | 0.16 | 0.08 | 0.64 | 0.00 | 0.27 | 0.98 |
| *12-month* | *pos* | -0.30 | 0.89 | 1 | -0.30 | 0.86 | 1 |
| | *neg* | -0.28 | 0.88 | 1 | -0.26 | 0.80 | 1 |

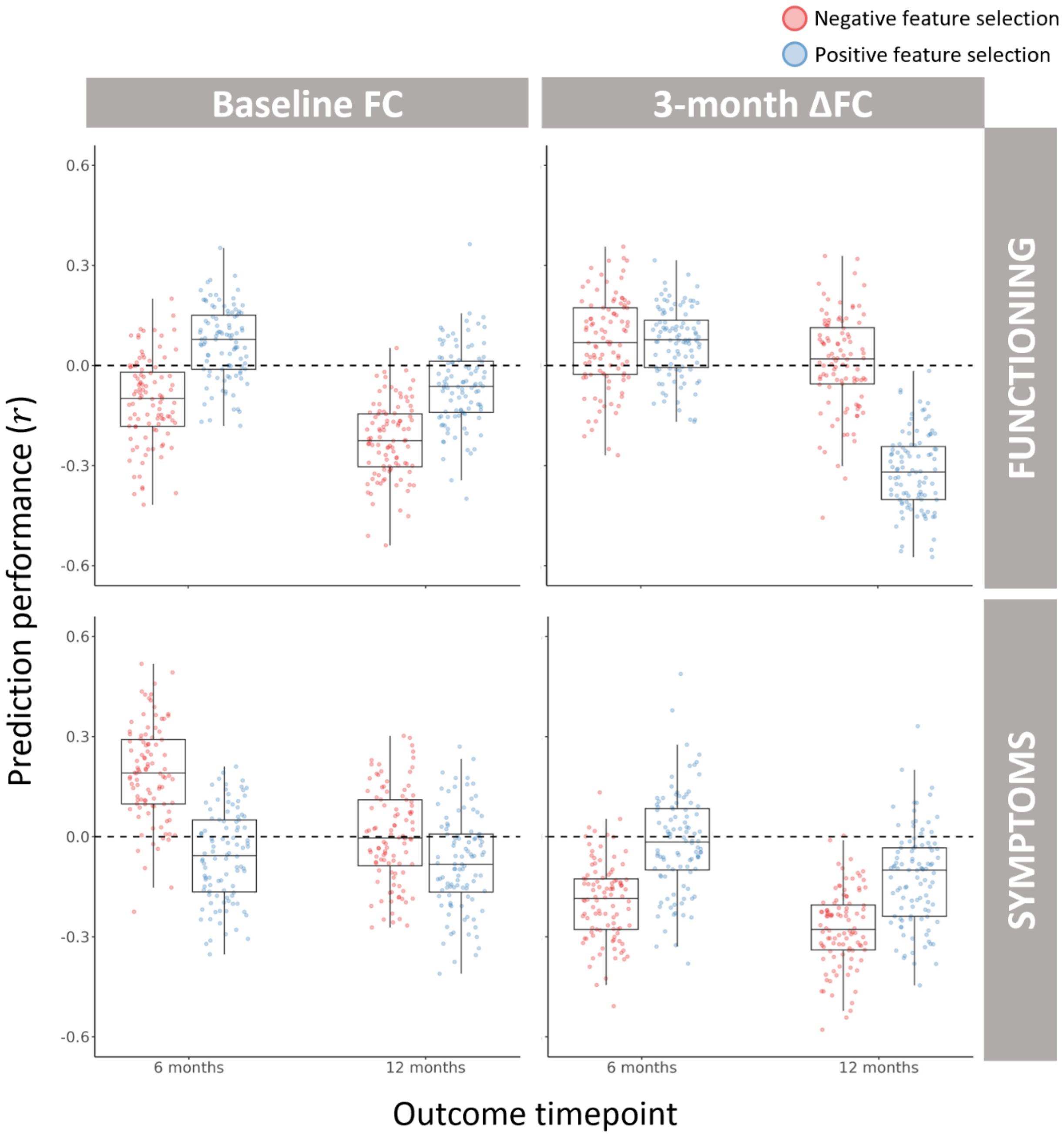


**Figure S8:** Performance of connectome-based predictive modelling (CPM) for predicting patients' clinical outcomes, using an alternate feature selection threshold of $p < 0.001$. Each data point shows the strength of Pearson's correlation between predicted and observed clinical outcomes for a single split of 4-fold cross-validation, with each of the 16 models comprising 100 random splits.

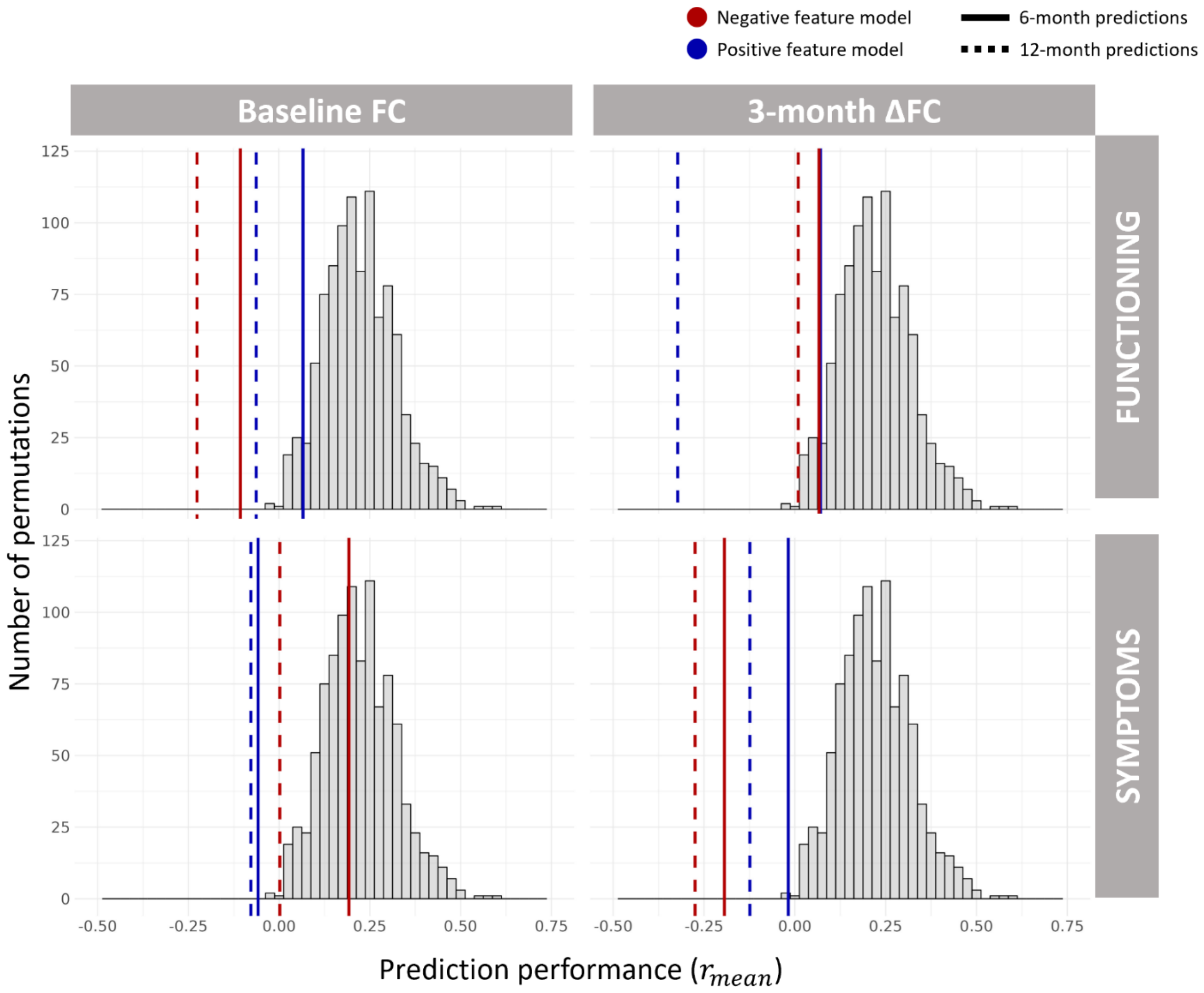


**Figure S9:** Prediction performance ($r_{mean}$ shown in red and blue) for all 16 connectome-based predictive modelling (CPM) models, with an alternate feature selection threshold of $p < 0.001$. Model performances are superimposed against a family-wise error (FWE)-corrected empirical null distribution derived by randomly permuting clinical outcomes amongst patients. For each of 1000 permutations, 100 splits of CPM were run to calculate a single null $r_{mean}$. FWE correction was applied across the 16 models shown.

**Table S3:** Prediction performance and significance for all 16 connectome-based predictive modelling (CPM) models, with an alternate feature selection threshold of $p < 0.001$. pos, positive feature model; neg, negative feature model.

| | | **Baseline FC** | | | **3-month ΔFC** | | |
|---|---|---|---|---|---|---|---|
| **SOFAS predictions** | | $r_{mean}$ | $p$ | $p_{FWE}$ | $r_{mean}$ | $p$ | $p_{FWE}$ |
| *6-month* | *pos* | 0.07 | 0.20 | 0.95 | 0.07 | 0.19 | 0.95 |
| | *neg* | -0.11 | 0.67 | 1 | 0.07 | 0.20 | 0.95 |
| *12-month* | *pos* | -0.06 | 0.51 | 1 | -0.32 | 0.97 | 1 |
| | *neg* | -0.23 | 0.91 | 1 | 0.01 | 0.31 | 1 |
| **BPRS predictions** | | $r_{mean}$ | $p$ | $p_{FWE}$ | $r_{mean}$ | $p$ | $p_{FWE}$ |
| *6-month* | *pos* | -0.06 | 0.50 | 1 | -0.02 | 0.34 | 1 |
| | *neg* | 0.19 | 0.06 | 0.60 | -0.19 | 0.80 | 1 |
| *12-month* | *pos* | -0.08 | 0.53 | 1 | -0.12 | 0.63 | 1 |
| | *neg* | 0.00 | 0.33 | 1 | -0.27 | 0.94 | 1 |
